# The Mathematics of Testing with Application to Prevalence of COVID-19

**DOI:** 10.1101/2020.05.31.20118703

**Authors:** Leonid Hanin

## Abstract

We formulate three basic assumptions that should ideally guide any well-designed COVID-19 prevalence study. We provide, on the basis of these assumptions alone, a full derivation of mathematical formulas required for statistical analysis of testing data. In particular, we express the disease prevalence in a population through those for its homogeneous subpopulations. Although some of these formulas are routinely employed in prevalence studies, the study design often contravenes the assumptions upon which these formulas vitally depend. We also designed a natural prevalence estimator from the testing data and studied some of its properties. The results are equally valid for diseases other than COVID-19 as well as in non-epidemiological settings.

## 1. Introduction

Uncovering the true scale of COVID-19 pandemic is critical for shaping public health policy and planning the comeback of economic and educational activities. This requires conducting large-scale testing studies for the ongoing and past disease aimed at determination of the prevalence of the disease in a given population. Knowledge of this parameter is also indispensable for estimating the true infection mortality rate of COVID-19.

A number of recently published studies of SARS-CoV-2 prevalence and COVID-19 sero-prevalence have stirred a considerable scientific and public controversy. The reasons include suboptimal study design and ritualistic use of not fully disclosed statistical methods that depend on various untestable assumptions. This creates an urgent need for methodological clarity regarding mathematical, statistical and epidemiological foundations of testing for COVID-19. The present article is a step in this direction. In particular, we explicitly state three fairly minimal, yet very essential assumptions underlying our analysis and discuss possible reasons for their violation using studies [1, 2] as an illustration. We provide detailed proofs of all the results that everyone can verify. The proofs depend on the three aforementioned assumptions alone, and we specify which assumptions are required for each of the results. Although it is uncommon for epidemiological studies to disclose all the details of statistical analysis, there is little doubt that some of our formulas were employed in published prevalence studies; however, the design of these studies often contravenes the assumptions upon which these formulas vitally depend.

In this article, we describe basic mathematical and statistical tools required for the analysis of data resulting from a population disease prevalence study. In particular, we compute in closed form (1) the distribution of the number, *n*, of recorded positive test results when testing *N* individuals using a test with sensitivity *α* and specificity *β*; (2) the conditional distribution of the number of true and false positive test results given that *n* positive test results were observed; and (3) the expected value and variance of the true number of infected individuals among those tested conditional on *n*, see Sections 3 and 4. We also prove in Section 5 an Equivalence Principle that extends all the above results to heterogeneous populations. Finally, we derive in Section 6 a natural statistical estimator of the disease prevalence and discuss some of its properties.

Based on our results, the prevalence of the current or past disease reported in a population study can be checked for self-consistency by comparing the observed number of positive cases with its theoretical prediction and by computing the expected number of false positive and false negative test results and all the associated probabilities, see Sections 3 and 4 for more details. Finally, the basic assumptions formulated in Section 2 may inform the design of future COVID-19 studies.

Given the current acute interest in SARS-CoV-2 and COVID-19, we will be using the terminology and biological considerations pertaining to COVID-19 testing. However, all the results are applicable in equal measure to testing for other diseases or conditions as well as in non-epidemiological settings.

The mathematical exposition in this article is accessible to biomedical scientists and health practitioners with a basic knowledge of discrete probability.

## 2. Basic Assumptions

We start by spelling out our assumptions. Their formulation and discussion are tailored to testing for COVID-19.

### A. Independence

*The events of a current or past infection in all tested individuals are stochastically independent*.

Although this assumption is indispensable for a valid statistical analysis, its violations in prevalence studies are common. One particular reason is excessive inclusion of members of the same household in a data set used for prevalence estimation. Among the aims of the Gangelt study [1], that was conducted on March 31-April 6, 2020 in the community of Gangelt in North Rhine-Westphalia, Germany, was to determine the excess risk of contracting COVID-19 for someone who lives in the same household with an infected individual. It was found that the risk of such secondary infection increases by 28.1%, 20.2% and 2.8% for households with two, three and four people, respectively, relative to the 15.5% risk of the primary infection [1]. Given that 919 participants of the Gangelt study for whom valid test results for SARS-CoV-2 and/or IgA, IgG antibodies were obtained belonged to only 405 households, the validity of Assumption A for this study is questionable. The same applies to the Santa Clara study, conducted on April 3-4, 2020 in Santa Clara county, CA [2]. In that study, among 3,390 participants whose specimens were analyzed there were 2,747 adults and 643 children living in the same household with some of these adults [2]. The validity of Assumption A is also problematic if disproportionately many study participants attended a known COVID-19 super-spreading event. In fact, investigation of the effects of one such event, carnival festival held around February 15, 2020 in Gangelt, revealed that the infection rate among the event participants was 2.6 higher than among non-participants and that the course of the disease was much more severe in the former than in the latter [1].

### B. Test Uniformity

*Both sensitivity and specificity of the test are identical for all tested individuals*.

Although almost universally accepted, this assumption may prove in ceratin cases questionable. For example, in some asymptomatic or pre-symptomatic carriers of COVID-19, the number of viral particles on a nasal or pharyngeal swab may be too low to be detectable by PCR test while the titer of IgA or IgM antibodies that serve as a marker of an ongoing disease may not exceed the detection threshold of a serological test. As yet another example, in convalescent COVID-19 patients the virus may already be undetectable while the titer of IgG antibody may be not detectable yet. In all such cases, the diagnostic sensitivity of the test will be reduced. Also, cross-reactivity with viral fragments or antibodies to another virus may increase the rate of false positive responses in those subjects who at the time of testing are, or have recently been, infected with another similar pathogen, e.g. a coronavirus causing the common cold. Such cross-reactivity would lead to a reduction in test specificity.

### C. Random Sampling

*N tested individuals were recruited by random sampling from the population of interest*.

The validity of this assumption may be thwarted by over-sampling from the same household. It may also prove suspect if recruitment for testing involves significant opportunity for self-selection making it likely that people who surmise that they have, or have had, the disease volunteer for the study. For example, the Santa Clara study recruited volunteers who responded to an advertisement posted on Facebook [2]. By contrast, the Gangelt study’s initial recruitment involved generating a random sample of 600 community members with distinct last names and mailing them an invitation to participate in the study, clearly in line with Assumption C. However, the research team allowed the participants to bring in other members of their household for testing. Out of these 600 individuals, 407 responded, which resulted in 1,007 study participants from 405 households who underwent testing for COVID-19 [1].

It is helpful to realize that the prevalence of present or past disease in a population may be quite heterogeneous across its various subpopulations. These subpopulations may be determined by such *observable* attributes as the presence of specific co-morbidities, age, sex, race, ethnicity, socioeconomic status and occupation, or their combinations. In the case of COVID-19, the effects of these attributes on the infection risk have been well-documented. For example, health care workers are generally exposed to much higher doses of SARS-CoV-2, and more frequently, than members of the general population and as a result have much higher prevalence of the disease. Other presently unknown variables associated e.g. with the genetic make-up or functioning of the immune and circulatory systems may prove important for proper prevalence stratification and estimation. Selection of subpopulations for a given study depends on the current knowledge about the epidemiology of the disease and the practicality of recruiting a sufficient number of study participants from each subpopulation.

In the Gangelt study, no explicitly defined subpopulations were specified except for those defined implicitly by the number of household members, co-morbidities and participation in the COVID-19 super-spreading event. In the Santa Clara study, the subpopulations were determined by sex, race and zip code. To mitigate significant violation of Assumption C (resulting, for example, in the fact that 63.7% of study participants for whom valid test results were obtained were females while the overall fraction of females in the Santa Clara county is 49.5%), the test results were subsequently re-weighted [2].

The overall logic and flow of exposition in the rest of the article is as follows. In Sections 3 and 4, we derive all our results, under Assumptions A and B, for a single homogeneous population where it can be assumed that the probability to have a current or past infection is the same for all individuals. Next, in Section 5, we establish, based on Assumption C, the *Equivalence Principle* for heterogeneous populations. This allows us extend all our results under Assumptions A-C to the general case of a heterogeneous population consisting of any number of homogeneous subpopulations. Finally, in Section 6 we use our results to design a prevalence estimator and discuss some of its properties.

## 3. Distribution of the Number of True and False Positive Test Results

Consider a test with a binary outcome (positive/negative) administered to *N* individuals. Let *α* be the sensitivity and *β* be the specificity of the test. Suppose that the test resulted in *n*, 0 ≤ *n* ≤ *N*, positive outcomes. Let *X, Y, Z* be the respective *unobservable* numbers of true positive, false positive and false negative test results and *U* = *X* + *Y* be the *observed* total number of positive results. Denote by *M* the unknown true number of presently or previously infected individuals (depending on the nature of the test) among *N* individuals selected from a population thought of, for the time being, as homogeneous with regard to prevalence of the disease. Below we seek to compute, under Assumptions A and B stated in Section 2, the distribution of random variables *X,Y, Z, U* and the conditional distributions of *X* and *Y* given that *U* = *n*. Additionally, in the next section we will compute in closed form the conditional expectation and variance of random variable *M* given that *U* = *n*.

It follows from Assumption A that the distribution of random variable *M* is binomial *B*(*N, p*), where *p* is the disease prevalence in the tested population:

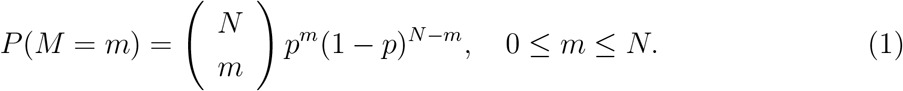

If *M* = *m* is fixed then each infected individual receives a positive test result, independently of other individuals (Assumption A), with the same probability *α*, the sensitivity of the test (Assumption B), or a negative test result with probability 1 − *α*. Then for the respective numbers, *X* and *Z*, of true positive and false negative test results we have

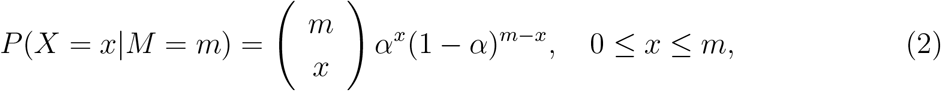

and

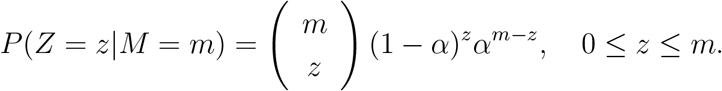

Similarly, according to Assumption B every healthy individual receives a false positive result with the same probability 1 − *β*, where *β* is the test’s specificity. Thus, for the random number, *Y*, of false positives we have

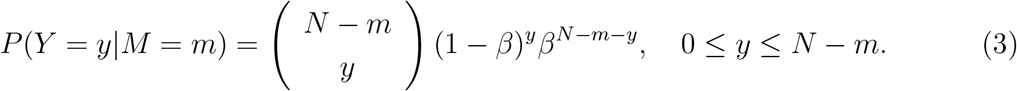

Importantly, it follows from Assumption A that, for every *m*, random variables *X, Y* and *Z* are conditionally independent given *M* = *m*.

Due to Assumptions A and B the number, *X*, of true positive test results is a *thinning* of the binomial random variable *M* with probability *α*. In general, thinning of a sequence of random events is their independent marking, or filtration, with the same probability; for more on thinning, see [3]. It is well-known, and easy to verify formally by compounding distributions (1) and (2), that *X* has binomial distribution *B*(*N, αp*). Likewise, *Y* is a thinning of the binomial random variable *N* − *M* with probability 1 − *β* and has the distribution *B*[*N*, (1 − *β*)(1 − *p*)] while the distribution of random variable *Z* is *B* [*N*, (1 − *α*)*p*].

We now compute the joint distribution of random variables *X* and *Y*. Notice that if *X* = *x* and *Y* = *y* then any admissible value, *m*, of random variable *M* must satisfy the inequalities *x ≤ m ≤ N* − *y*. Using the formula of total probability, invoking equations (1)- (3), rearranging the factors, making a change of variable *j* = *m* − *x* and finally employing Newton’s binomial formula we have for all *x, y ≥* 0 such that 0 ≤ *x* + *y* ≤ *N*

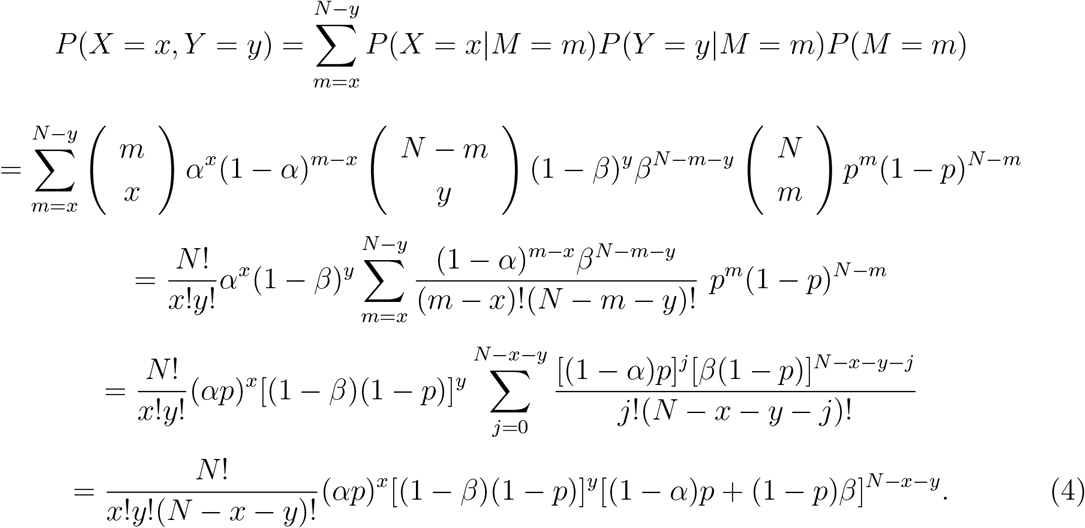

Thus, random vector (*X*, *Y, N* − *X* − *Y*) has trinomial distribution

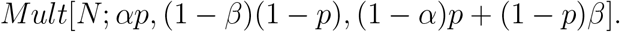

From this we conclude by lumping together true and false positive test results and combining the probabilities of these outcomes that the distribution of *U* = *X* + *Y*, the total number of positive test results, is binomial *B*(*N, λ*), i.e.

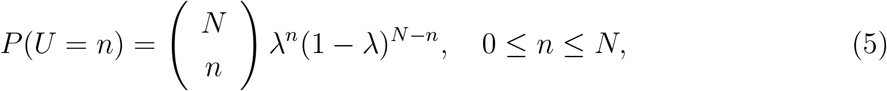

where

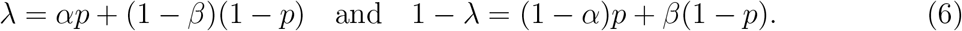

Formulas (4) and (5) lead to the following conditional distribution of the number of true and false positives given the observed total number of positive results:

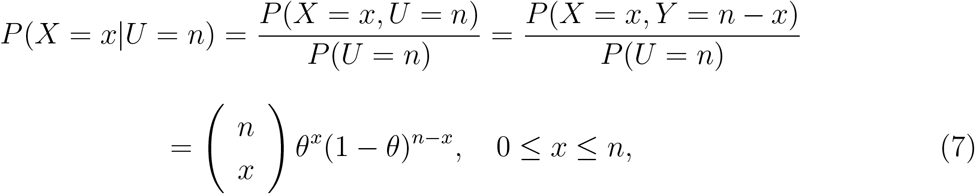

and

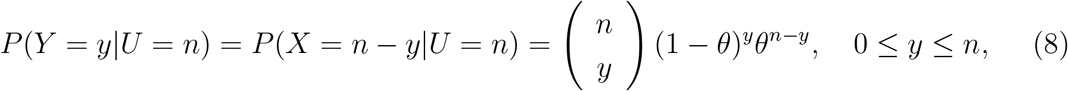

where

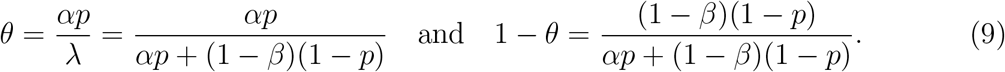

Therefore, the posterior distributions of the number of true and false positives given the observed number, *n*, of positive test results are *B*(*n, θ*) and *B*(*n*, 1 − *θ*), respectively.

Formula (7) is remarkably similar to the one describing the conditional distribution *P*(*X* = *x|X* + *Y* = *n*), 0 ≤ *x ≤ n*, in the case of two independent Poisson random variables *X* and *Y* [4, Section 9.3, pp. 277-278].

Distributions (7) and (8) have three notable features:

a. They are independent of the number, *N*, of tested individuals;
b. Parameters *θ* and 1 − *θ* given by (9) represent the *predictive positive and negative values* that can be obtained by applying the Bayes formula to the prior probabilities 1 − *p* and *p*, respectively;
c. Distributions (7) and (8) depend on a *single* parameter

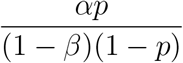

that combines the basic parameters *p, α, β*.

The extraordinary simplicity of formulas (5), (7) and (8) should not becloud the fact that their validity depends critically on Assumptions A and B.

## 4. The Expected Number of Infected Individuals for a Given Number of Positive Test Results

As will be shown in Section 6, population prevalence can be estimated based on a formula for the conditional expectation of the number, *M*, of infected individuals given *U* = *n* positive test results. The conditional distribution of random variable *M* given that *U* = *n* has the following form

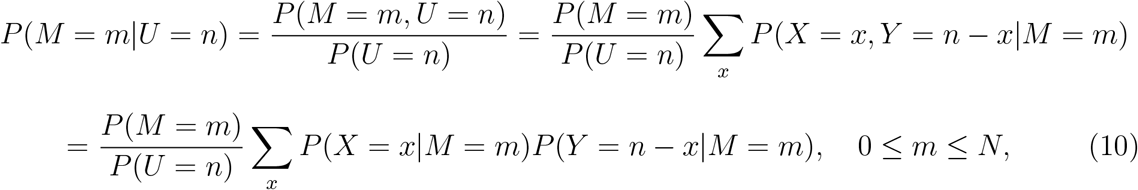

where *P*(*U* = *n*) was specified in (5)-(6) and *x* must satisfy the inequalities 0 ≤ *x* ≤ *m, x* ≤ *n* and *n* − *x* ≤ *N* − *m* or equivalently

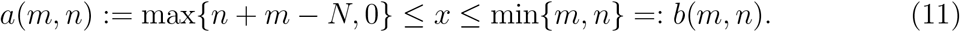

Although formulas (1)-(3) and (5) combined with (10) lead to an explicit expression for the conditional probability *P*(*M* = *m*|*U* = *n*), the latter does not seem to be reducible to a simple form. However, the corresponding expectation and variance can be computed in closed form, as we show below.

According to (10), in order to find the conditional expectation 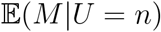 and variance *Var*(*M|U* = *n*), we need to compute, for *ℓ* = 1, 2, the following quantity:

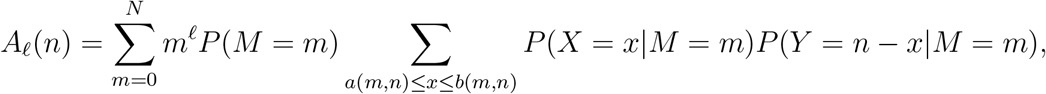

where the bounds for variable *x* are given in (11). The range for pairs (*x*, *m*) has a simpler form

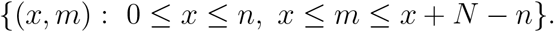

Therefore, switching the order of summation, changing the variable in the internal sum to *j* = *m* − *x* and using formulas (1)-(3) we obtain

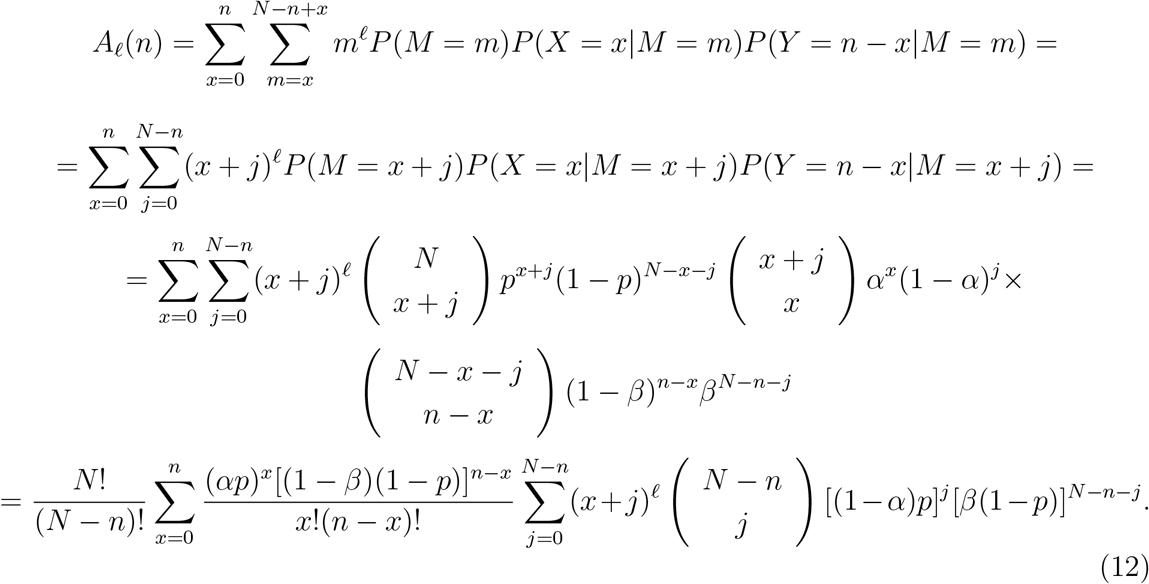

Using (6) we represent the internal sum in the last formula for *ℓ* =1 as

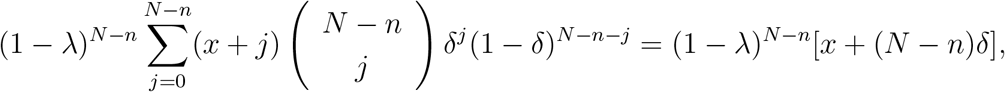

where we set *δ* = (1 − *α*)*p*/ (1 − *λ*) and employed the formula for the expected value of the binomial distribution *B*(*N − n,δ*). We now continue the above derivation of the formula for *A*_1_(*n*) as follows:

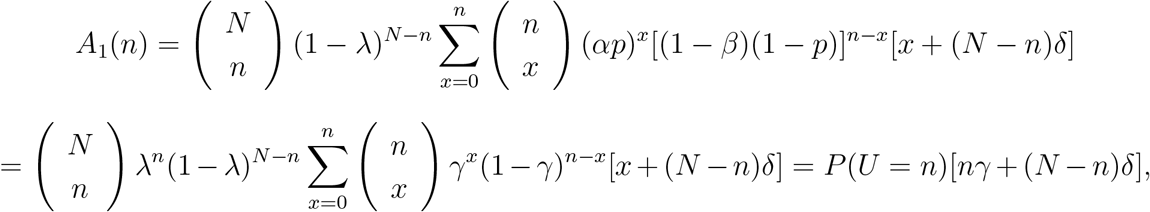

where we set *γ* = *αp/λ* and used (5) along with the formula for the expected value of the binomial distribution *B*(*n*, *γ*). Thus, from (10) we obtain

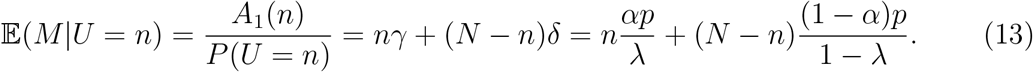

Similarly, in the case *ℓ* = 2 we use formulas for the expected value and second moment of the distribution *B*(*N − n, δ*) to represent the internal sum in (12) in the form

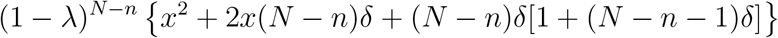

which yields

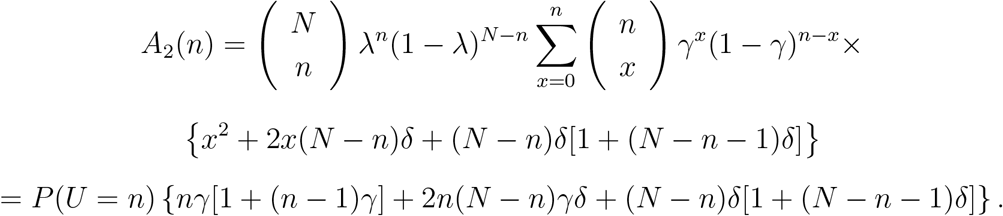

Therefore, from (13) and (6) we finally obtain

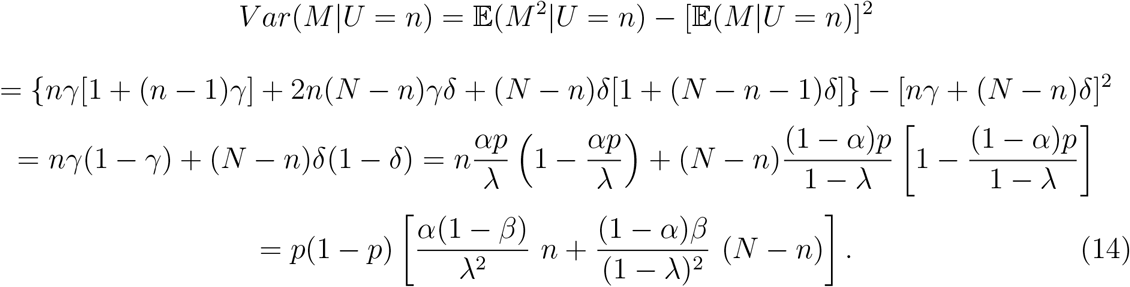

Inspection of formulas (13) and (14) shows that the computed conditional expectation and variance depend only on the following two combinations of parameters *p, α, β*:

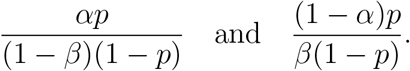

## 5. The Equivalence Principle for Heterogeneous Populations

In Sections 3 and 4 we considered a population that was assumed homogeneous in the sense that all individuals in the population had the same probability, *p*, to have a current or past infection. The goal of this section is to show how all our results can be generalized to a heterogeneous population consisting of *r* homogeneous subpopulations. Let **w** = (*w*_1_*, w_2_, …,w_r_*), where 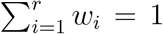, be the vector of relative sizes (weights) of these subpopulations and **p** = (*p*_1_*, p*_2_, *…, p_r_*) be the vector of their disease prevalences.

For a non-negative integer vector **x** with *r* components, we will use the following notation: 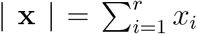 and 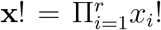. Also, for two such vectors **x** and **y** we denote 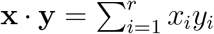, **xy** = (*x*_1_*y*_1_, *x*_2_*y*_2_, *…, x_r_y_r_*) and 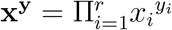. Finally, **y** ≤ **x** means that *y_i_ ≤ x_i_* for all *i* =1, 2, …, *r*.

Let *N_i_*, 1 ≤ *i* ≤ *r*, be the number of individuals from *i*-th subpopulation among *N* tested individuals. Then under the random sampling assumption (Assumption C), random vector **N** = (*N*_1_, *N*_2_,…, *N_r_*) with *|***N***|* = *N* has multinomial distribution *Mult*(*N*, **w**):

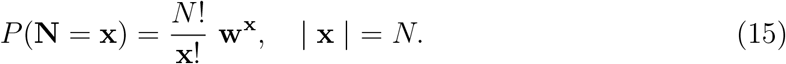

Denote by *M_i_* the number of infected individuals among *N_i_*, 1 ≤ *i* ≤ *r*. Random vector **M** = (*M*_1_, *M*_2_,…, *M_r_*) represents a component-wise thinning of random vector **N** with the vector of thinning probabilities **p**. What is the distribution of random vector **M**? The computation given below shows that, by contrast to the binomial case, it is not multinomial!

Assumption A and subpopulation homogeneity imply that that for any *i*, 1 ≤ *i* ≤ *r*, the distribution of random variable *M_i_* conditional on *N_i_* = *x_i_* is binomial *B*(*x_i_, p_i_*). Also, conditional on **N** = **x**, where |**x**| = *N*, components of random vector **M** are, according to Assumption A, independent. Therefore, for any vector **y** such that |**y**| ≤ *N*, we have using (15) and the formula of total probability, making change of variable change **z** = x − **y** and finally using the multinomial formula

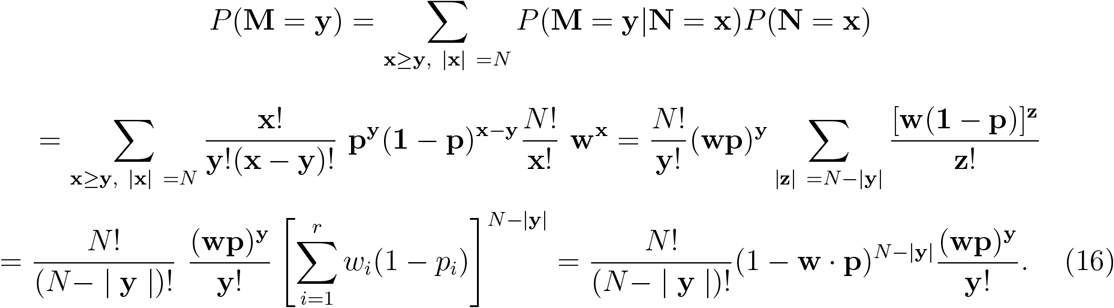

Due to Assumption B all computations made in Sections 3 and 4 involve only the total number, *m* = *|***M***|*, of infected individuals among those tested. Using the multinomial formula we derive from (16) the distribution of random variable *|***M***|*:

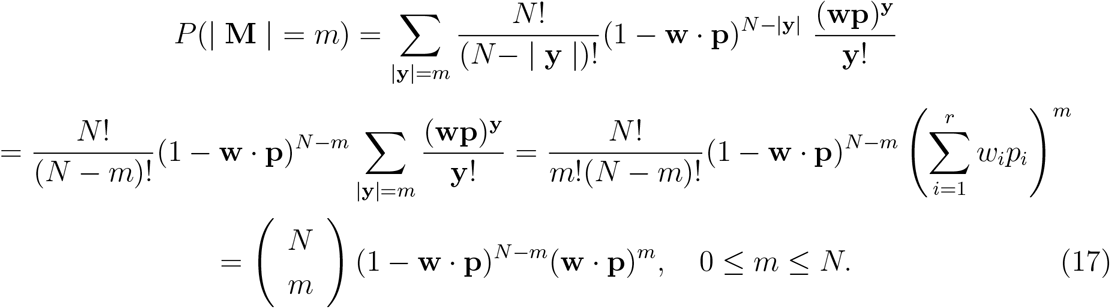

Therefore, the total number of infected individuals among *N* people tested follows binomial distribution *B*(*N*, **p** · **w**). Comparison between formulas (17) and (1) leads to the following **Equivalence Principle**:

*The distribution of the number of infected individuals among N individuals selected at random from a heterogeneous population consisting of r homogeneous subpopulations with weights w*_1_, *w*_2_, …, *w_r_ and disease prevalences p*_1_, *p*_2_, *…, p_r_ is the same as for a homogeneous population with prevalence*

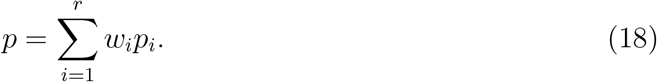

Thus, all the results in Sections 3 and 4 derived for a homogeneous population with disease prevalence *p* are valid for a heterogeneous population if one selects *p* in accordance with formula (18). This conclusion is, of course, only natural; however, it depends on Assumptions A-C in very essential ways.

## 6. Prevalence Estimation

If *N* individuals randomly drawn from a population of interest were tested and *n* positive test results were observed then a “naïve” estimate of the prevalence of the current or past disease in the population is *p*_0_ = *n/N*. The testing process can be thought of as the following mental experiment: for an infected individual, a coin is flipped that lands “heads” with probability *α* and “tails” with probability 1 − *α* while for a healthy individual, another coin is flipped that lands “heads” with probability 1 − *β* and “tails” with probability *β*. In these terms, *p*_0_ is the fraction of “heads” recorded for *N* independent replications of this random experiment. Therefore, *p*_0_ depends on the sensitivity and specificity of the test and the prevalence of the disease. To untangle them and find an estimator of the prevalence alone, recall that the distribution of the number of positive test results is binomial *B*(*N, λ*), see Section 3. Because *p*_0_ is a consistent unbiased estimator of *λ* = *αp* + (1 − *β*)(1 − *p*) we can set for the desired estimate, 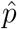, of the population prevalence *p* the following equation

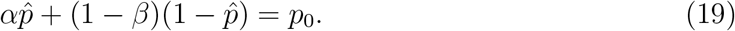

Thus, 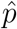 is the population prevalence that would produce, on average, the same fraction of positive test results when *N* individuals are tested as the one actually observed. Solving this equation and assuming that *α* + *β >* 1 (a condition always met in practice) we obtain

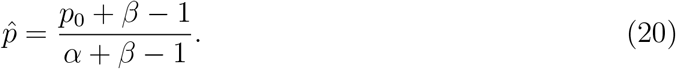

Note that this estimator was employed in [2]. Observe that 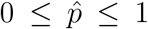 if and only if 1 − *β < p*_0_ *< α*. Thus, estimator *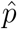* is not feasible in the cases where population prevalence is smaller than the test’s false positive rate or larger than the test’s specificity.

Estimator (20) can be obtained alternatively on the basis of the conditional distribution of the number, *M*, of infected individuals among those tested given the observed number of positive test results. Recall that in view of (13)

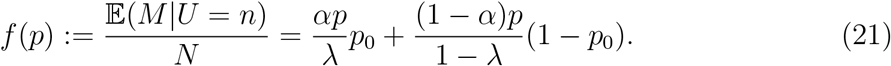

The main difficulty here is that this estimator depends on the unknown prevalence parameter *p* that we want to estimate. A natural idea, then, would be to find the value of *p* for which *f* (*p*) = *p* and take it as a prevalence estimator. Using expression (6) for λ we get after some algebra

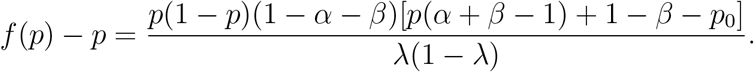

Therefore, in view of (20) in the case where *α* + *β >* 1, 0 *< p <* 1 and 1 − *β ≤ p*_0_ *≤ α* the required fixed point of function *f* is exactly the above estimator 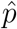! Therefore, according to (21) the estimator 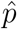 represents the expected fraction of infected individuals among those tested given the observed number of positive test results.

The estimator 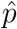 has the following remarkable feature that can be termed the “mixture-invariance” property. Suppose that the population of interest consists of *r* subpopulations, each of which is either homogeneous or a mixture of homogeneous sub-subpopulations. Let *N_i_* ≥ 1 be the number of tested individuals from *i*-th subpopulation and *n_i_* be the number of positive outcomes, based on the same test. Denote by 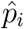 the above prevalence estimate for *i*−th subpopulation and set 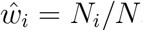, where *N* = *N*_1_ + *N*_2_ + … + *N_r_*. Finally, assume that *α* + *β >* 1 and 1 − *β ≤ n_i_/N_i_ ≤ α* for all *i*. Then for *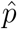*, the prevalence estimate for the entire population, we have

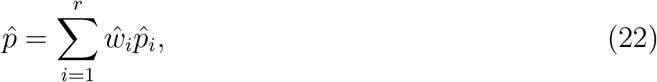

compare with (18).

In fact, using formula (20) we get

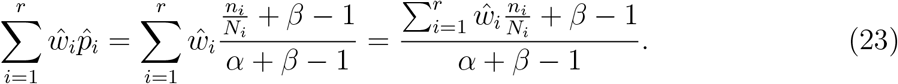

Observe that

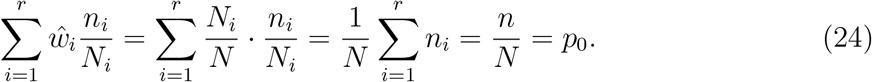

It follows from (24) that *p*_0_ is a weighted average of the ratios *n_i_/N_i_*, which implies, due to their above-assumed bounds for these ratios, that 1 − *β ≤ p*_0_ *≤ α*. We now continue (23) to find that

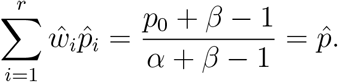

The invariance property (22) can be used to combine prevalence estimates for several subpopulations of a given population obtained within the same study, or different studies utilizing the same test, into a prevalence estimate for the entire population.

## 7. Discussion and Recommendations

In this article, we derived formulas for the distribution of the number of positive test results for a test with given sensitivity and specificity administered to *N* individuals randomly selected from a given population as well as for the conditional distribution of the unobservable number of true and false positive test results given the observed number, *n*, of positive test outcomes. Also, we found closed form expressions for the expected value and variance of the unknown true number of infected individuals among those tested conditional on the observed value of *n*. These results led us to a natural estimator, 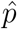, of the population prevalence, see formula (20). We initially derived our results in Sections 3 and 4 for a homogeneous population; however, in Section 5 we gave a rigorous proof of the *Equivalence Principle* stating that in the case of a heterogeneous population, one has only to replace the population prevalence with the average of the prevalences of homogeneous subpopulations of the given population weighted by their relative sizes, see formula (18). Finally, we proved a similar “mixing-invariance” property for the prevalence estimator, see formula (22).

Importantly, we spelled out the three assumptions required for the validity of each of these formulas explicitly, see Section 2. We discussed these assumptions and illustrated possible reasons for their violation using the well-known Gangelt [1] and Santa Clara [2] studies as examples. It is somewhere between likely and certain that some of the formulas derived in this article were used in one form or another in the published COVID-19 prevalence studies. However, it is also certain that suboptimal study design of some of these studies violated the assumptions upon which these formulas critically depend. This contradiction may cast doubt on the validity of COVID-19 prevalence estimates that resulted from these studies.

Note that in this article we never used any asymptotic arguments, i.e. those applicable to large values of *N*. Therefore, our results can be used for any sample size *N*, no matter how small.

We conclude with a list of specific recommendations regarding the design of testing studies and statistical methodology informed by our analysis.

1. Including in the same prevalence analysis of multiple groups of subjects from the same household and disproportionately large groups of individuals who participated in the same COVID-19 super-spreading event should be avoided. The same applies to individuals who were in close and/or protracted contact with each other without PPE. Likewise, self-selection of subjects for the study makes prevalence estimates unreliable.
2. Knowledge disease prevalence estimates, resulting from the data obtained on the same testing platform, for subpopulations with known demographic weights leads automatically, through the aforementioned “mixing-invariance” principle, to a prevalence estimate for a heterogeneous population comprised of these subpopulations without the need for a separate *de novo* study.
3. The “naïve” prevalence estimator *p*_0_ = *n/N* is a function of the true population prevalence and the test’s sensitivity and specificity. The correct “disentangled” estimator, 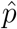, is given by formula (20). Additionally, *p*_0_ may deviate considerably from 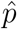. For example, for a perfectly specific test (*β* = 1), one has 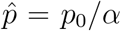. Thus, the use of *p*_0_ as prevalence estimator should be discouraged.
4. Prevalence estimation for a population with high prevalence requires a very sensitive tests while for populations with low prevalence, a very specific tests should be employed.

## Data Availability

All the data is provided in the manuscript itself.

